# Identifying COPD subtypes using multi-trait genetics

**DOI:** 10.1101/2023.02.20.23286186

**Authors:** Andrey Ziyatdinov, Brian D. Hobbs, Samir Kanaan-Izquierdo, Matthew Moll, Phuwanat Sakornsakolpat, Nick Shrine, Jing Chen, Kijoung Song, Russell P. Bowler, Peter J. Castaldi, Martin D. Tobin, Peter Kraft, Edwin K. Silverman, Hanna Julienne, Hugues Aschard, Michael H. Cho

## Abstract

Chronic Obstructive Pulmonary Disease (COPD) has a simple physiological diagnostic criterion but a wide range of clinical characteristics. The mechanisms underlying this variability in COPD phenotypes are unclear. To investigate the potential contribution of genetic variants to phenotypic heterogeneity, we examined the association of genome-wide associated lung function, COPD, and asthma variants with other phenotypes using phenome-wide association results derived in the UK Biobank. Our clustering analysis of the variants-phenotypes association matrix identified three clusters of genetic variants with different effects on white blood cell counts, height, and body mass index (BMI). To assess the potential clinical and molecular effects of these groups of variants, we investigated the association between cluster-specific genetic risk scores and phenotypes in the COPDGene cohort. We observed differences in steroid use, BMI, lymphocyte counts, chronic bronchitis, and differential gene and protein expression across the three genetic risk scores. Our results suggest that multi-phenotype analysis of obstructive lung disease-related risk variants may identify genetically driven phenotypic patterns in COPD.

## Introduction

Chronic Obstructive Pulmonary Disease (COPD) is a phenotypically heterogeneous disease^1^. Some have hypothesized that COPD is a syndrome constituted of multiple disease subtypes involving different biological mechanisms^2,3^. Understanding the molecular basis underlying this heterogeneity in COPD can advance our knowledge of COPD etiology and improve patient treatment. However, efforts in learning specific disease mechanisms and potential COPD subtypes using molecular markers have been hampered by multiple issues, including variability in measurement across time and conditions, variability across targeted tissues, and reverse causation (e.g., when the trait is influenced by disease treatment).

Germline genetic variants are present from birth and do not vary with time, disease, or treatment, and thus offer a relevant alternative for learning subtypes. Moreover, genome-wide association studies (GWAS) have now been conducted on a vast number of human traits and diseases, providing an increasingly precise overview of shared genetic effects across human phenotypes (pleiotropy). Building on these two features, several methods have been proposed for inferring genetically driven disease subtypes.^4-7^ Broadly speaking, these methods leverage the relationships between disease-associated variants and other phenotypes to construct clusters of variants based on similarity in their multitrait association pattern. The variants within each group can be further characterized and might ultimately be used to classify individuals. COPD is a strong candidate for such disease subtype inference, as we and others have demonstrated that genetic variants associated with COPD have substantial pleiotropic effects^1,8^.

In this work, we examined genetic variants identified in GWAS of moderate-to-severe COPD; spirometry, as COPD is defined by decrements in lung function; and asthma, an obstructive lung disease with extensive clinical and physiological overlap with COPD. We applied a Bayesian method previously used in type 2 diabetes, another complex disease characterized by disease heterogeneity^5^, to cluster these variants based on their association with a broad range of traits measured in the UK Biobank, a large population cohort with hundreds of phenotypic measures available. We then used individual-level genetic and phenotypic data from the COPDGene^9^ study, a cross-sectional cohort of COPD cases and controls with deep pulmonary phenotyping and extensive clinical data, to investigate potential subtypes based on the identified clusters.

## Methods

### Selection of genetic variants associated with COPD, lung function, and asthma

Genetic variants relevant to COPD were identified from three obstructive lung disease-related GWAS^1,8,10^: COPD itself^1^, spirometric lung function phenotypes^8^ and asthma^10,11^. We relied on published results from the largest genome-wide association studies for the first two datasets. For asthma, we conducted a custom meta-analysis of public GWAS asthma results from UK Biobank^11^ and the GABRIEL consortium^10^. Our list included 164, 279, and 45 variants for COPD, lung function, and asthma, respectively (see **Supplementary Methods**), and a total of 482 variants after removing six duplicated variants that were identical between COPD and lung function. For the primary analysis of clustered variants, as the causal variants were not known, we retained some variants in linkage disequilibrium with the top GWAS variants. For analyses that required independent variants, we performed an additional LD-pruning, removing variants correlated across the four subsets (e.g., variants selected in the asthma GWAS that might be correlated with variants selected from the COPD GWAS) using *SNPclip* (ldlink.nci.nih.gov/?tab=snpclip) and an r^2^ threshold of 0.1 in 1000 Genomes European ancestry subjects, and resulting in 377 independent variants.

### Selection and analysis of traits from UK Biobank

We built an agnostic strategy to select traits from 2,409 GWAS summary statistics of quantitative and binary traits derived in the UK Biobank (expanding on prior work)^8^ to be used in the cluster’s inference. We applied multiple filters to remove non-informative and low-quality GWAS from the collection. First, we excluded traits directly related to lung function, COPD, and asthma (e.g., respiratory diagnosis codes and different measures of asthma). Second, we removed attributes with low effective sample size (N_eff_ < 200,000) to ensure that the included GWASs carried a similar amount of information. Third, we removed traits that did not show any genome-wide significant association (P < 5×10^−8^) for at least one of the 482 selected variants. Fourth, we removed GWAS displaying high pairwise Pearson correlations of Z-scores, as implemented in the R package caret^12^ (r^2^ threshold = 0.8). Finally, we oriented all Z-scores to COPD risk-increasing alleles and divided Z-scores by the square root of the effective sample size, thus, converting Z-scores to standardized effect sizes. As an additional requirement for implementing the clustering approach (see next section), we split each row of the Z-score matrix into two meta-trait Z-scores, positive and negative. That resulted in doubling the number of traits in the downstream clustering analysis.

### Clustering by Nonnegative Matrix Factorization

The clustering of variants was performed using the Nonnegative Matrix Factorization (NMF) method, which has shown promising results in disease subtype learning^5^. In brief, NMF decomposes the 2*T* × *S* matrix of Z-scores, where *T* denotes the number of traits, and *S* the number of variants, into a lower-dimensional representation *Z* ≈ *WH*^*T*^ with weight matrices *W* and *H* of sizes 2*T* × *K* and *S* × *K*, respectively, and where *K* is a latent dimension to be learned from the data. More specifically, we applied a probabilistic Bayesian model of NMF^13^ that iteratively learns the weights in *W* and *H*. The approach includes four key steps: (i) reducing a reconstruction error through the *β*-divergence function; (ii) adding a *K*_*λ*_-length vector of relevance weights *λ* as an auxiliary variable (*K*_*λ*_ = 32, by default); (iii) using half-normal priors on weights (L2-norm regularization); and (iv) satisfying the non-negative constraints on weights (*W* > 0, *H* > 0). Altogether, this Bayesian implementation of NMF automatically learns the latent dimensionality *K* and avoids ambiguity compared to other NMF algorithms^13^. In practice, the number of learned dimensions *K* is obtained by taking non-zero entries in the *K*_*λ*_-length of relevance weights *λ*.

### Building cluster-specific Genetic Risk Scores

Weighted genetic risk scores (GRS) were derived for each of the *K* clusters inferred from the NMF using the variants weight from the matrix *H*. More specifically, for a cluster *i* and variants *g*_*j*_ with *j* = *1* … *S*, the weighted cluster-specific GRS are calculated as *GRS*_*i*_ = ∑_*S*_ *H*_*ji*_*g*_*j*_. For comparison purposes, we also defined a baseline GRS defined as the unweighted sum of genotypes: *GRS*_*0*_ = ∑_*S*_ *g*_*j*_. Note that because the *H*_*j*=1…*K*_ does not sum to 1, the sum of the *GRS*_*i*_ is not equal to *GRS*_*0*_.

### COPDGene dataset

The characteristics of various genetic risk scores were examined using individual-level data from the COPDGene study. The COPDGene study (NCT00608764, www.copdgene.org) recruited 10,198 non-Hispanic white (NHW) or African-American (AA) participants, aged 45-80 years old, with at least 10 pack-years of smoking and no diagnosed lung disease other than COPD or asthma^9^. IRB approval was obtained at all study centers, and all study participants provided written informed consent. Illumina (San Diego, CA) performed genotyping on the *HumanOmniExpress* array, and imputation to HRC 1.1 was performed using the Michigan Imputation Server. COPDGene subjects were extensively phenotyped, with data collected using questionnaires, spirometry, and inspiratory and expiratory CT scans at baseline. Subjects were invited to participate in follow-up visits, including spirometry and CT scans, and a subset had cell counts and biomarkers^14^. In this study, we considered a total of 240 traits (**Supplementary Table S2**). All traits were tested for association with the cluster-specific GRS described in the previous section. All models were adjusted for relevant covariates such as age, sex, pack-years of smoking and smoking status, scanner and center as appropriate.

### Statistical tests and models fit in COPDGene

We fit linear and logistic regression models for 240 quantitative and binary traits respectively modeling the effect of the four genetic risk scores (*GRS*_0…3_) using individual-level genetic and phenotypic data from the COPDGene cohort. We first estimated the marginal effect of each *GRS*_*i*_ using a standard univariate model: *f*(*Y*)∼*GRS*_*i*_ + *cov*, where *cov* represents trait specific covariates. We then assessed the impact of decomposing *GRS*_0_ into the *GRS*_1…3_ using two approaches. First, we compared the above marginal model against a conditional model including *GRS*_0_: *Y*∼*GRS*_0_ + *GRS*_*i*_ + *cov* using a likelihood ratio test (LRT). Second, we assessed the overall contribution of all three *GRS*_1…3_ by comparing the marginal model for *GRS*_0_ against a joint model, including all three cluster-specific *GRS*_*i*_ : *Y*∼*cov* + *GRS*_0_ + ∑_*i*=1…3_ *GRS*_*i*_, again using a LRT. This test of heterogeneity, referred to further as *P*_het_, quantifies the improvement in model fitting when decomposing *GRS*_0_ into *K GRS*_*i*_. To examine the relative contribution of the *GRS*_1…3_ to heterogeneity, we also extracted effect estimate from this joint model. For individual GRS significance derived from the marginal, conditional, and joint model, we use the stringent Bonferroni correction threshold of 7 × 10^−5^, accounting for 723 tests conducted for each model. For *P*_het_, we applied a Benjamini and Hochberg^15^ correction and reported results with False Discovery Rate (FDR) <0.1. Note that both Bonferroni and FDR correction can be conservative as they do not account for the correlation between the traits tested.

### Association of GRS with gene expression, protein biomarkers, and clinical outcomes in COPDGene

We tested whether cluster-specific GRSs were associated with changes in gene expression using RNA-sequencing from peripheral blood taken at the 5-year follow-up visit in COPDGene. We used limma/voom^16^, and adjusted all analyses for age, sex, smoking status, ancestry principal components, and batch. We performed a similar analysis with SomaScan plasma proteomics data, adjusting for age, sex, smoking status, and ancestry principal components. Besides investigating associations of the GRSs with phenotypes and omics data in COPDGene, we also sought to determine whether the cluster-specific GRS could identify individuals with higher or lower risk of lung-related phenotypes. We used the top and bottom decile of subjects based on high scores on *GRS*_1_, and low scores on *GRS*_2_and *GRS*_3_, and first examined the association of eosinophils and steroid use, adjusting for age, sex, ancestry-based principal components, and GOLD stage. Finally, we investigated heterogeneity in medication use conditional on the same percentile of *GRSs*, using information on COPD exacerbations and medication treatments available in COPDGene (see Supplemental Table 7).

## Results

### Overview of the multi-trait genetic approach

To address our objective of identifying potentially distinct COPD subtypes, we implemented a three-step workflow. First, we identified a list of 482 genetic variants associated at genome-wide significance level with COPD, lung function (FEV_1_ and FEV_1_/FVC), or asthma (**Table S1**). For each of these variants, we assembled and harmonized a matrix of association Z-scores for a broad range of traits derived from the UK Biobank. Second, we derived a lower-dimensional matrix representation using the non-negative matrix factorization (NMF) algorithm, followed by an *ad hoc* thresholding of the variants assignment weights, resulting in the stratification of variants into non-overlapping clusters potentially representing different genetic pathways of lung function. Third, we built a genetic risk score (GRS) for each of the inferred clusters, and investigated the link between these GRSs and COPD-related phenotypes using individual-level data from COPDGene participants (**Figure 1**).

**Figure 1.**
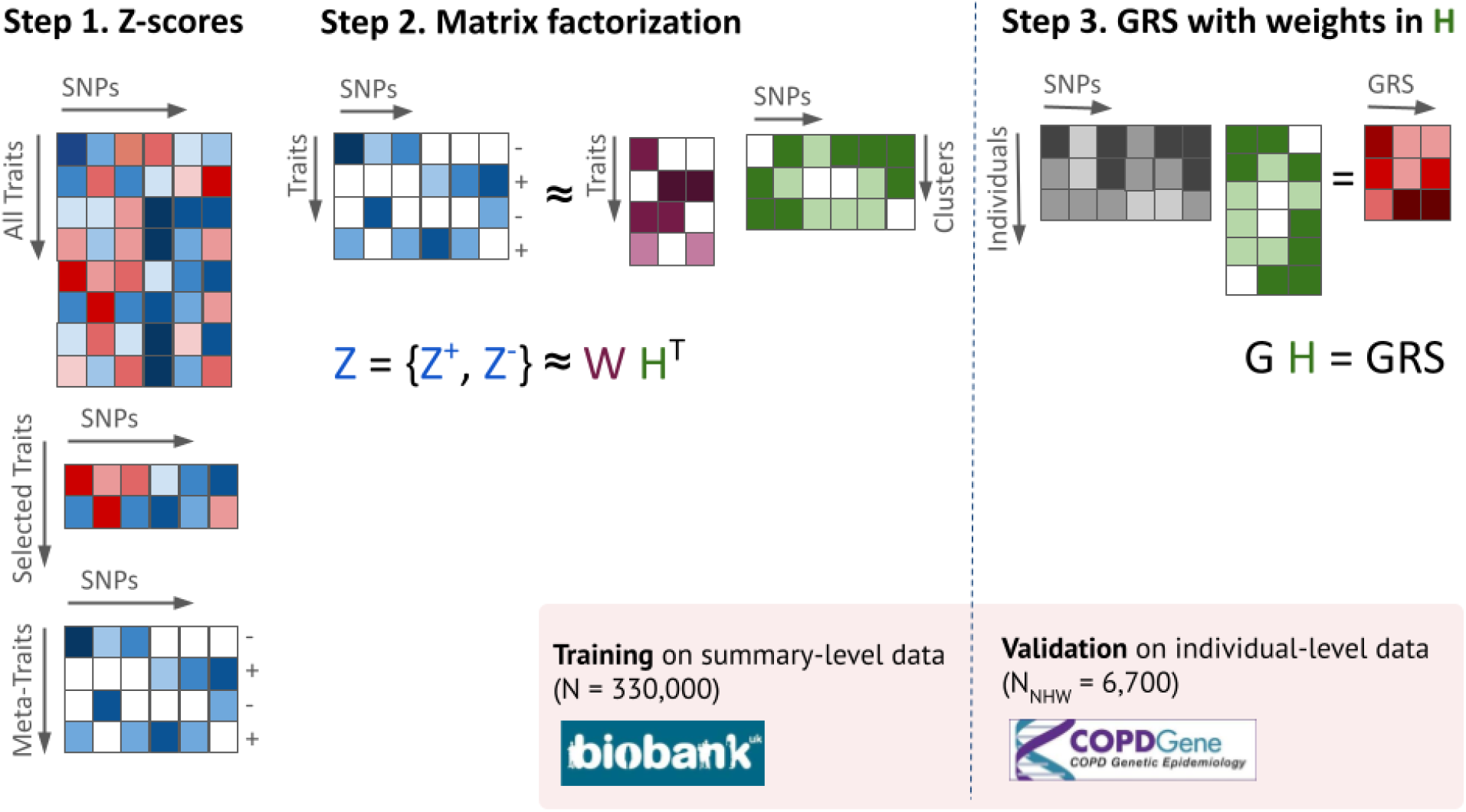
Analysis pipeline for COPD subtypes inference using multi-trait genetic approach. At step 1, the multitrait Z-score matrix derived from UK Biobank data is thinned to a smaller set of traits driven by pre-selected variants. Each selected trait is further split into two meta-traits with either positive or negative Z-scores. At step 2, the matrix of processed Z-scores is decomposed into products of two weight matrices W and H, with the number of columns K being equal to the number of clusters. Finally at step 3, the weight matrix for variants, H, is used to build K weighted GRSs at the individual level data from validation cohort (COPDGene).

### Variant characteristics and cluster inference

We first cross-examined the COPD genetic association with association at the three other phenotypes (COPD, asthma, FEV_1_ and FEV_1_/FVC), using a subset of 377 independent variants out of the 482. A total of 52%, 80%, and 61% variants genome-wide significant with asthma, FEV_1,_ and FEV_1_/FVC, respectively, were nominally significant (P < 0.05) in the COPD GWAS. All of those nominally significant variants were also directionally consistent. Quantitatively, Z-scores for COPD were significantly correlated with asthma (*ρ* = 0.44, P = 3.1 × 10^−10^), FEV_1_ (*ρ* = -0.81, P=1.4 × 10^−90^), and FEV_1_/FVC (*ρ* = -0.88, P=2.5 × 10^−123^) (**Fig. S1**). Interestingly, asthma and COPD display a mixture of two distributions, suggesting the presence of two genetic mechanisms with a different contribution to the two outcomes (**Fig. S1a**). We then extracted the association between the larger set of 482 variants and traits measured in the UK Biobank cohort. We filtered and harmonized a total of 2,409 UK Biobank phenotypes, selecting outcomes displaying evidence of association with variants, removing phenotypes with low sample sizes, and removing redundancy due to high phenotypic correlation (see **Methods**). After this processing, the Z-score matrix included 44 phenotypes with association Z-score for all 482 variants. No clear pattern emerged from a visual inspection of this matrix when using a standard hierarchical clustering algorithm (**Fig. S2**).

However, the application of the NMF algorithm to this matrix identified three clusters with associated weight matrices *W* and *H*, reflecting the contribution of traits and genetic variants, respectively. To characterize cluster compositions in trait dimension, we operated on normalized trait weights (unit sum of cluster weights in columns of *W*) and identified the top normalized trait weights for each cluster (**Fig. 2**). We oriented all weights such that positive and negative weights of normalized traits reflect increasing and decreasing risk of COPD, respectively. Cluster 1 displayed high positive weights for wheeze, eosinophil percentage, and neutrophil counts. Cluster 2 had negative weights for traits linked to body composition and obesity (hip circumference, body fat, and BMI). Cluster 3 displayed positive weights for height, grip strength, and birth weight, and negative weights for blood cell counts.

**Figure 2.**
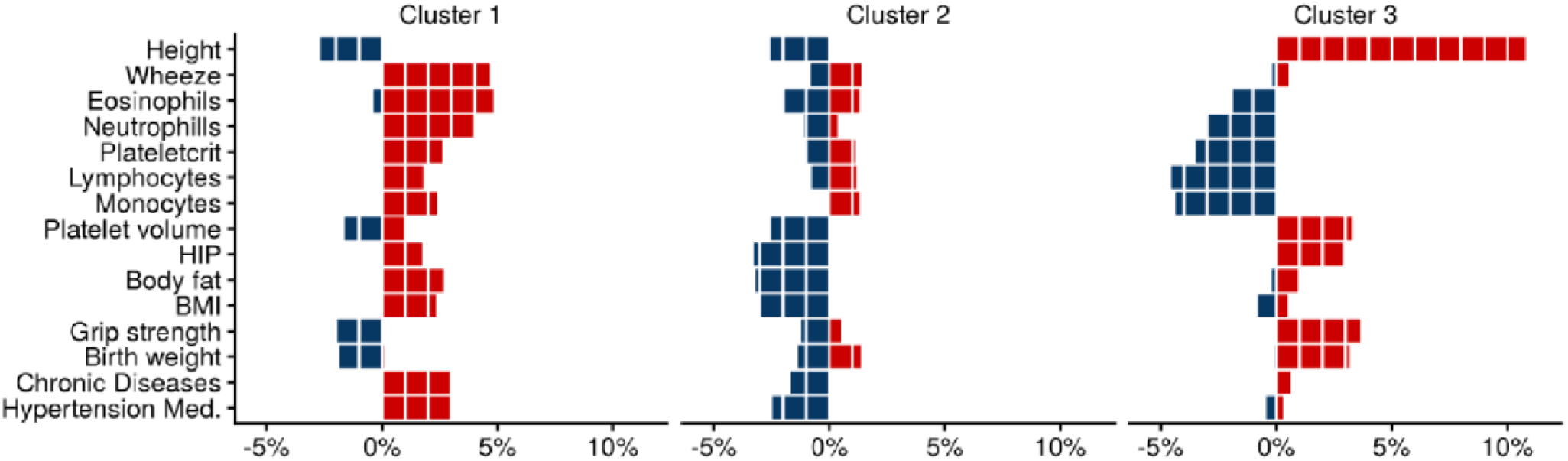
Distribution of trait weights across the three inferred clusters. We selected the top 15 traits with the largest contribution to the cluster inference. The Nonnegative Matrix Factorization (NMF) clustering constructs two matrices W and H out of the Z-score association matrix, so that Z≈WH^T^, where H is a matrix of traits weight with number of columns equals to the number of clusters. The top traits corresponded to those harboring normalized weights (unit sum of column elements) larger than 3% for at least one cluster. The figure represents the weights for each trait and each of the three inferred clusters. Red bars correspond to the contribution of positive Z-scores submatrix, and blue bars to negative Z-scores submatrix.

To characterize cluster compositions by variants, we derived variant weights (unit sum of variant weights in rows of H) and assigned variants with weights >50% to the corresponding cluster. Approximately 78% of all variants match that criterion, with 156, 148, and 78 variants selected for clusters 1, 2, and 3, respectively. The assignment of variants across clusters is illustrated in the alluvial plot in **Figure S3**. We compared the origin of these variants –*i*.*e*. whether they were selected from COPD, lung function or asthma GWAS– against the expected from a random assignment (out of the 482 variants, 34%, 57%, and 10% were selected from the COPD, lung function, and asthma GWAS, respectively). Cluster 1 variants display a small enrichment for asthma variants and a reduced representation of lung function variants (33%, 51%, and 16% variants from the COPD, lung function, and asthma sets, respectively). The overrepresentation of asthma variants in this cluster is consistent with the composition of traits (**Figure 2**), where wheeze and eosinophil percentage have the largest weights. Conversely, in cluster 2, lung function variants were slightly overrepresented and asthma variants underrepresented (34%, 64%, and 2% variants from the COPD, lung function and asthma sets, respectively). Cluster 3 did not display specific enrichment (35% 59%, and 8% variants from the COPD, lung function and asthma sets, respectively).

### Clinical features of inferred clusters of variants in COPDGene

To determine whether the inferred clusters of variants were related to COPD phenotypes, we constructed three cluster-specific weighted genetic risk scores (*GRS*_*1*_, *GRS*_*2*_ and *GRS*_*3*_), and an unweighted genetic risk score (*GRS*_*0*_) including all 482 variants, that we applied to individual-level genotypes from COPDGene (Methods, **Fig. 1**). We first tested the marginal association between each of the four GRSs (*GRS*_0_, *GRS*_*1*_, *GRS*_*2*_ and *GRS*_*3*_) and 240 features and outcomes measured in COPDGene (**Table S2**). As expected, *GRS*_0_, which include all variants, displayed the strongest (by Z-score) association with most phenotypes and was close to the best association from *GRS*_1. .3_ (**Fig. S4, Table S3**), with the exception of height peak expiratory flow (PEF), and steroid treatment. **Figure 3** presents the effect estimates and standard errors of all four GRS for selected outcomes representing different phenotypic groups (lung function, anthropometric measurements, imaging, etc). Focusing on nominally significant signals, *GRS*_*1*_ was associated with the highest eosinophils, highest self-reported steroid treatment, and the lowest six-minute walk distance. Cluster 2 was associated with the lowest FEV_1_/FVC ratio and highest emphysema fraction. Cluster 3 was associated with higher height and FEV_1_/FVC, and the largest risk of coronary disease.

**Figure 3.**
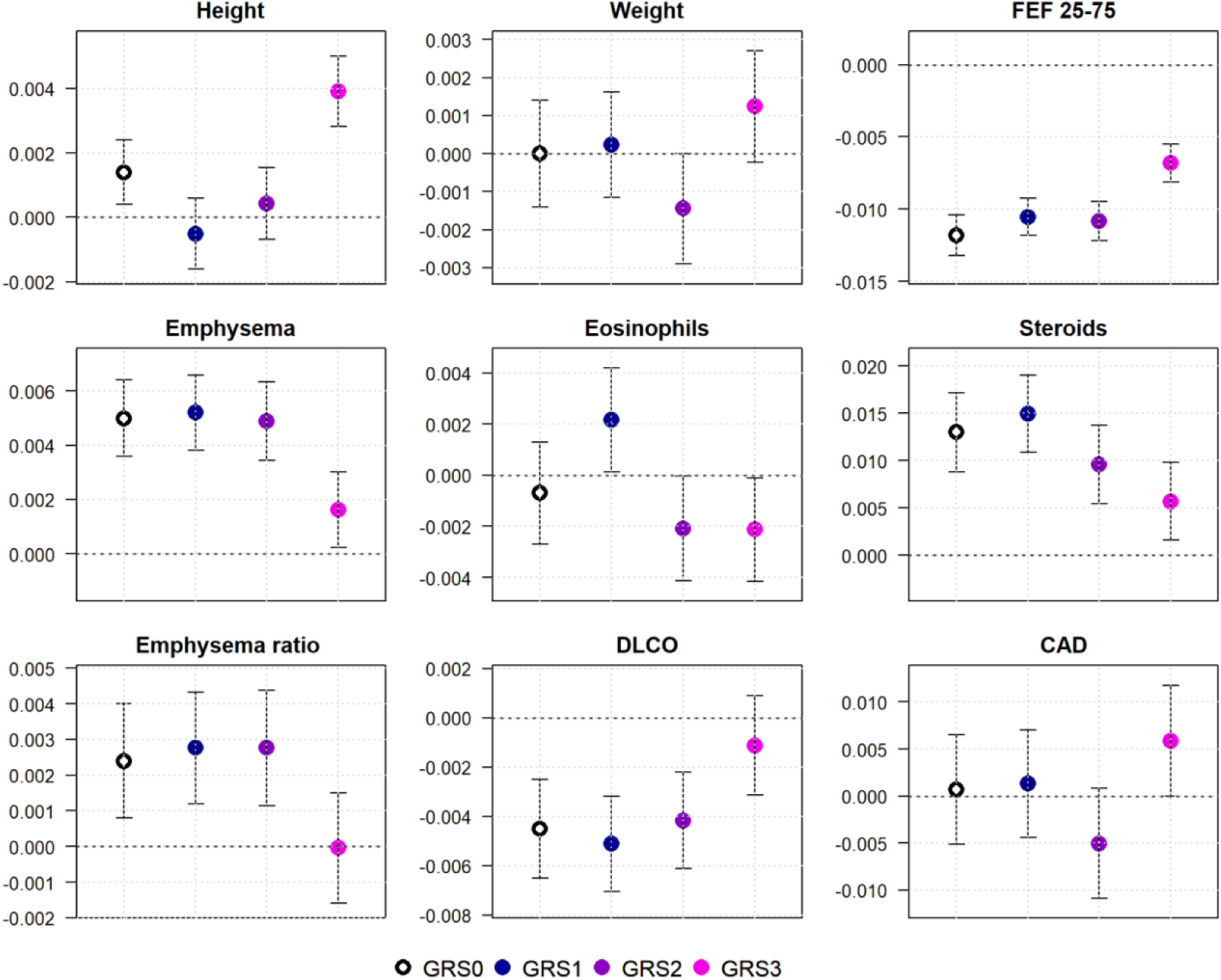
Marginal effects of GRSs on selected traits in the validation COPDGene dataset. Point estimates and 95% confidence intervals obtained from marginal models are displayed for cluster-specific GRSs (***GRS***_**1**…**3**_; from dark blue to pink) and unweighted GRS (***GRS***_**0**_; black). For comparison purposes, all GRS were re-scaled to a unit variance. We selected traits representing different COPD phenotypic groups: height, weight, forced expiratory flow at 25–75% of force vital capacity (FEF25-75), visual emphysema score (Emphysema), eosinophils count (Eosinophils), steroids treatment (Steroids), upper third/lower third emphysema ratio (Emphysema ratio), diffusing capacity for carbon monoxide (DLCO), coronary artery disease (CAD).

For each trait, we then performed a test of heterogeneity comparing a joint model including all four *GRS*_*0*…3_ against a baseline model including only *GRS*_*0*_. Overall, 47 traits out of 240 showed a significant effect of including cluster-specific *GRS*_*1*_, *GRS*_*2*_ and *GRS*_*3*_ in addition to *GRS*_*0*_ (column P_het_, **Table S3**). **Figure 4** presents the relative contribution of the three GRSs of this conditional model for these 47 traits. *GRS*_*1*_ and *GRS*_*2*_ were significant for most phenotypes, suggesting a complementary contribution. We also found many of the same trait associations as in the UK Biobank. *GRS*_1_ showed enrichment for significant association with blood cell counts and inflammatory biomarkers, including C-Reactive Protein and Selectin, and hepatocyte growth factor (HGF / c-MET), previously implicated in COPD pathogenesis. *GRS*_2_ showed association with obesity and related traits including BMI, insulin, coronary-artery disease, and sleep apnea. Finally, *GRS*_*3*_ showed association with height, most clinical lung function measurements, and COPD-related phenotypes including CC16, a biomarker with previous associations with COPD. Altogether these results offer an indirect validation of the trait weights learned in the UK Biobank dataset (**Figure 2**) and suggest that the derived partitioned GRS can partly capture heterogeneity of clinical features related to COPD.

**Figure 4.**
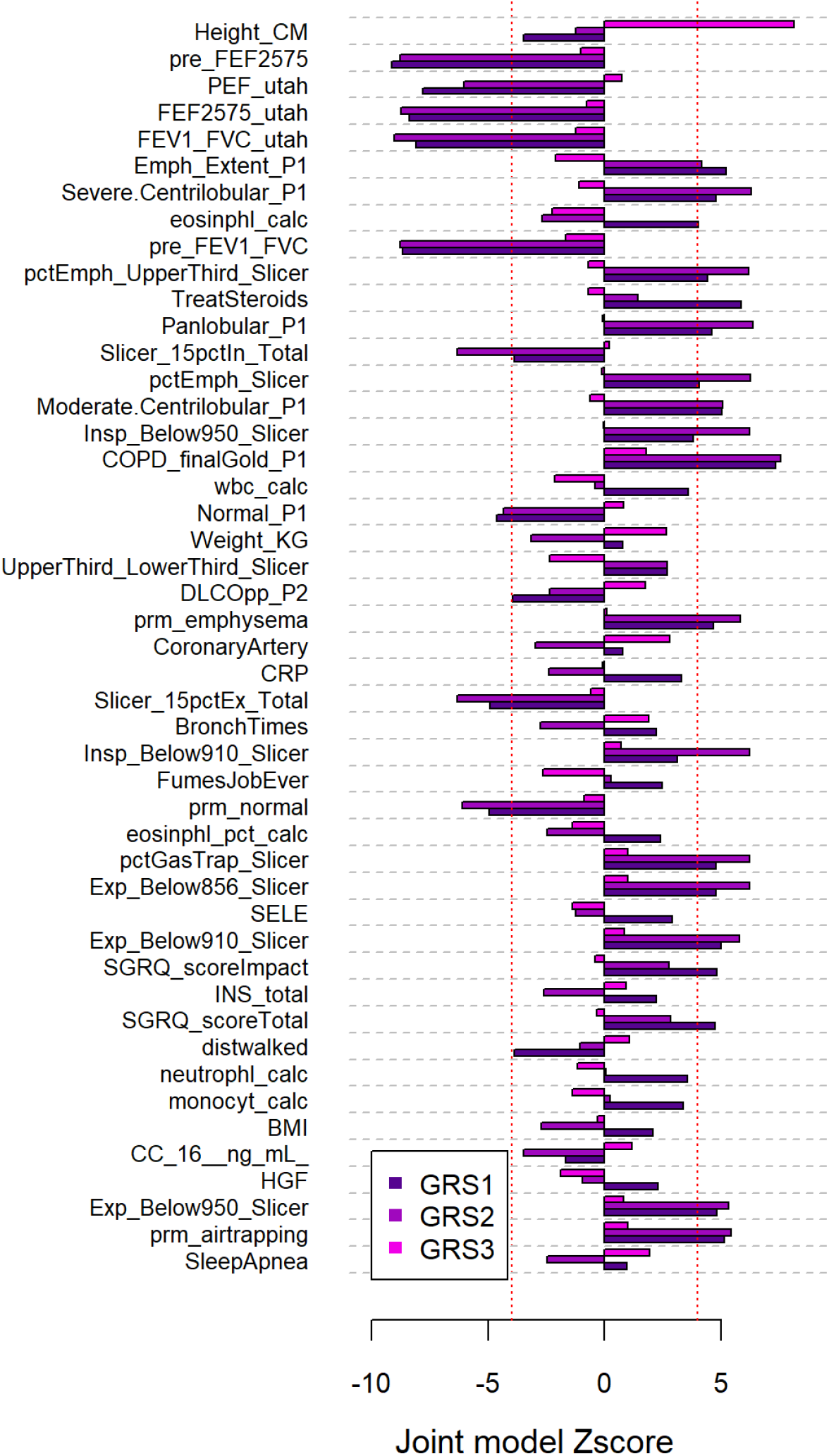
Contribution of cluster-specific GRSs. Out of 240 COPDgene phenotypes tested for association with genetic risk scores, a total of 47 phenotypes showed a statistically significant (*P*_het_) improvement of model fit at an FDR of 0.1 when comparing the marginal GRS_0_ model against a full model including GRS_0_ and all GRS_1-3_. The barplots represent the relative contribution of GRS_1_, GRS_2_, and GRS_3_, measured as Zscore derived from the full model, for these 47 phenotypes, highlighting which of the three GRS convey the improved fit. Phenotypes are order by *P*_het_. Red dash lines indicate the stringent Bonferoni significance threshold accounting for a total of 723 tests.

### Functional enrichment and GRS-stratified participants characteristics

We first conducted in silico functional enrichment analysis for variants within each cluster using FUMA^17^ and a limited set of annotations (**Table S4** and **Supplementary Methods**). Cluster 1 showed a significant enrichment for immunity and inflammation pathways that was consistent across multiple input databases, and in agreement with all previous results. Cluster 2 harbored significant enrichment for a single annotation related to endocytosis. Cluster 3 showed enrichment for a broad range of pathways covering many biological components and cellular processes.

We then tested the association of the GRSs with peripheral blood RNA-Sequencing and SomaScan proteomics data using individual-level data from COPDGene. Data were available on 2,666 subjects for gene expression, and 3,687 subjects (4,979 protein levels) for SomaScan proteomics^18^. At an FDR of 0.1, we found 1, 2, and 18 differentially expressed protein-coding for *GRS*_1_, *GRS*_2_, and *GRS*_3_, and a total of 7 differentially expressed proteins. Top results are shown in **Supplemental Tables S5**-**S6**. *GRS*_1_ was associated with ANKRD35. GRS2 with NISCH and ILF3; while the top results for *GRS*_2_ in protein were not significant, results included IL-17 RC and SDF1 (stromal derived factor-1). *GRS*_3_ was associated with multiple genes in the MHC region on chromosome 6 including ZFP57, BTN3A2, HLA-A, H4C13, HLA-DQB1, and HLA-DQB2 as well BT3A3, MICA, MICB, and C4B. Protein results also included FTMT, a mitochondrial ferroxidase enzyme, and RGAP1 (encoded by *RACGAP1*), neither in the MHC region.

Finally, we explored whether cluster-specific GRS could identify individuals with higher or lower risk of specific clinical outcomes. Based on the results of heterogeneity testing, we examined high scores on *GRS*_1_, and low scores on *GRS*_2_ and *GRS*_3_, and tested the association with eosinophils. The top decile had a significantly higher level of eosinophils, after adjusting for age, sex, ancestry-based principal components, and GOLD stage (*P* = 0.007), and a 1.66 (95% CI, 1.10-2.52) fold increased odds of reporting requiring steroid treatment. We also investigated potential heterogeneity in treatment among COPDGene participants harboring extreme values of the genetic risk scores (**Figure S5)**. When comparing the characteristics of the top and bottom 5^th^ percentiles of the three GRSs, we observed strong heterogeneity in corticosteroid, steroid, and theophylline treatments when stratifying participants by *GRS*_1_. Groups defined by *GRS*_2_ were marked by differences in long-acting beta-antagonist and ipratropium treatments.

## Discussion

COPD is characterized by a simple and effective diagnostic criterion based on spirometry. This diagnosis has arguably led to greater recognition of the disease, effective bronchodilator therapy, and improved outcomes. However, patients with COPD demonstrate substantial clinical heterogeneity. Identifying the molecular basis for this heterogeneity has proven challenging. One major challenge to explaining COPD heterogeneity is the long course of the disease. Most phenotypic characteristics, such as exacerbations, blood cell counts, and degree of emphysema, are affected not only by severity but also by disease course and effects of treatment. Assessing COPD heterogeneity using genetic variants offers an opportunity to assess clinical heterogeneity without these confounders.

In this work, we explored a multi-trait genetic approach based on a set of genetic variants associated with COPD, lung function, and asthma. We identified three different groups of genetic variants. Although these variants were taken from three sources (COPD, lung function, and asthma), variants did not simply segregate by their source. This finding is particularly notable for asthma, for which genetic risk appears to be enriched for immune cells and overlap with autoimmune disease, in contrast to COPD and lung function loci, for which genetic signals are enriched in regulatory regions from lung tissue^1,8^.

These results were further supported by the analysis in COPDGene. COPDGene data was not used for the phenotypic association for clustering, and thus the consistent associations of height, body mass index, and cell counts confirm these association results. These three groups of genetic variants included one related to eosinophils and inflammatory biomarkers, a second related to lower body mass and greater emphysema; and a third with higher height, the strongest associations with lung function, and an association with coronary disease. The individual GRSs demonstrated associations with gene expression and protein biomarkers. While aside from the association of GRS_3_ with the HLA region (likely driven by genetic variants in this region), most of these associations were relatively weak, they do support the hypothesis that these GRSs reflect differing biology. Variants comprising *GRS*_1_ appear to affect *ANKRD35*, a paralog of *ZDHHC13* that may be associated with granulocyte count^19^. For *GRS*_2_, *Nisch* modified mice exhibit an emphysema-like phenotype^20^ and *ILF3* is reduced in BAL from COPD patients compared with never smoking controls^21^. For *GRS*_3_, in addition to multiple HLA associations, FTMT is involved in ferroptosis, known to be an important pathway in COPD^22^, and RGAP1 (in addition to MICB) was recently in a Mendelian Randomization analysis of proteomics with lung function^23^.

Our results also suggest that GRSs may be able to identify patients that may benefit from more specific treatments. Using a combination of GRSs consistent with higher eosinophil burden based on UK Biobank phenotypes, we identified subsets of COPDGene participants with different eosinophil counts and history of steroid use. Our method avoids some of the confounding present in phenotypic and/or other molecular data. Genetic variants are stable biomarkers and can be used for prediction prior to the development of disease or during disease treatment. Our ability to identify clusters and associations was based on the largest genetic association studies available to date across multiple phenotypes in the UK Biobank. However, even with 482 variants to predict relevant subtypes or disease axes^2^, power is likely still limited^24^. Several of these signals likely represent the same causal signal, and many associated phenotypes in the UK Biobank were highly correlated, reducing the number of analyzed traits, and the UK Biobank lacks many of the respiratory phenotypes that may be useful for COPD phenotypes (such as imaging). Nevertheless, our analysis based on GWAS summary statistics is versatile and can be replicated in new coming datasets of larger sample sizes and new phenotypes, including imaging and molecular endotypes, particularly if relevant to respiratory traits. Future work in genetic subtyping could also allow for the inclusion of additional sub-genome-wide significant variants (as done in current polygenic risk scores) and functional genetics, including cell types and mechanisms, all of which should further improve our ability to identify heterogeneity from genetic profiles.

In summary, we clustered genetic variants associated with obstructive lung disease using a diverse set of phenotypes, identifying three multi-trait/multi-variant disease scores. These scores demonstrate different associations with biologic and clinical phenotypes, which promise to improve as GWAS and other omics studies expand.

## Data Availability

All data produced in the present work are contained in the manuscript.

## Web resources

https://pheweb.org/UKB-SAIGE/pheno/495: UK Biobank GWAS results for asthma^11^ (meta-analyzed with asthma GWAS results from the GABRIEL consortium^10^)

## Acknowledgements

MHC was supported by R01HL149861, R01HL135142, R01HL137927, R01HL147148, and R01HL089856. This work was supported by NHLBI U01 HL089897 and U01 HL089856. The COPDGene study (NCT00608764) is also supported by the COPD Foundation through contributions made to an Industry Advisory Committee comprised of AstraZeneca, Bayer Pharmaceuticals, Boehringer-Ingelheim, Genentech, GlaxoSmithKline, Novartis, Pfizer and Sunovion. M.H.C. has received grant support from GSK and Bayer, consulting or speaking fees from Genentech, AstraZeneca, and Illumina. EKS has received grant support from GSK and Bayer.

The content is solely the responsibility of the authors and does not necessarily represent the official views of the NIH. The funding body has no role in the design of the study and collection, analysis, and interpretation of data and in writing the manuscript.

This research was conducted using the UK Biobank Resource under Application #20915. This research has been conducted as part of the INCEPTION program (Investissement d’Avenir grant ANR-16-CONV-0005). This research was supported by the Agence National pour la Recherche (ANR-20-CE36-0009-02).

MDT is supported by Wellcome Trust Awards WT202849/Z/16/Z and WT225221/Z/22/Z. The research was partially supported by the National Institute for Health Research (NIHR) Leicester Biomedical Research Centre; the views expressed are those of the author(s) and not necessarily those of the National Health Service (NHS), the NIHR or the Department of Health. This research was funded in part, by the Wellcome Trust. For the purpose of open access, the author has applied a CC BY public copyright license to any Author Accepted Manuscript version arising from this submission.

