## Supplemental Material for "Identifying COPD subtypes using multi-trait genetics"

### Supplementary Material

### Supplementary Methods

#### Independent associations from public GWAS

To select lung disease-associated variants for clustering analysis, we mined results from public GWAS of COPD[1], lung function phenotypes[2] and asthma[3,4]. Methods to identify independent associations varied across studies. The COPD GWAS[1] revealed 82 independent genome-wide significant loci ($P < 5 \times 10^{-8}$). Secondary signals were then obtained by conditional and joint analysis at each of the 82 loci, adding other 82 secondary variants and resulting in the total number of 164 variants for COPD. The GWAS of four lung function phenotypes[2], forced expired volume in 1 second (FEV1), forced vital capacity (FVC), FEV1/FVC and peak expiratory flow (PEF), reported 139 independent genome-wide significant loci ($P < 5 \times 10^{-9}$) associated at least with one phenotype. Further follow-up joint analysis of the 139 loci identified 140 additional variants, resulting in a total of 279 lung function variants. Our custom meta-analysis of two asthma GWAS from UK Biobank[4] and the GABRIEL consortium[3] yielded 45 independent variants. It was not enriched for secondary signal at those loci.

#### Enrichment analysis with FUMA

We used FUMA[5] SNP2GENE function (https://fuma.ctglab.nl/snp2gene) to associate SNPs with protein coding genes based on two criteria, the physical position (in 30kb radius of a gene) and eQTLs (all significant cis-eQTL from GTEx up to a distance of 1Mb). For the latter, we restrained the eQTLs to the one that were found in relevant tissue for the trait under study (COPD, Asthma, lung function), including immune cells, blood vessels, lung, and adipocyte. Note that the FUMA SNP2GENE function requires association *p*-values. Here we used the minimum *p*-value reported across the three studied traits. For genes mapped within each clusters, we then performed a functional enrichment for pathways defined in KEGG[6], GO[7], BioCarta, Reactome[8], databases, and the ad hoc *immunologic_signature* gene sets, using the FUMA GENE2FUNC function. For the enrichment *p*-values, the cluster's gene were compared against a background of protein coding genes.

#### Supplementary Figures

**Figure S1. Distribution of Z-scores for asthma, FEV1 and FEV1/FVC as a function of COPD Z-score across the 377 independent variants**

Association Z-score for COPD across 377 independent variants plotted as a function of association Z-score for asthma (a), FEV1 (b) and FEV1/FVC (c). Variants selected from the COPD GWAS are indicated by a black circle. Variants nominally significant with COPD are indicated in orange.

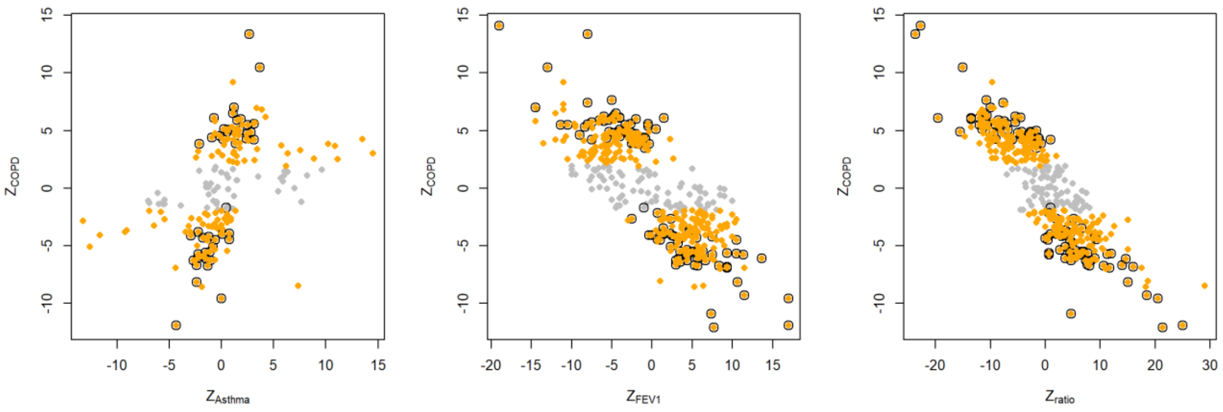

**Figure S2. Heatmap of the Z-scores matrix for the 44 traits and 482 variants**

The color encodes the gradient of association Z-scores from positive (blue) to negative (red) between the 482 variants and the 44 traits. Rows and columns were ordered using a hierarchical clustering based on Ward.D2 methods applied to Euclidean distance. The resulting clustering of SNPs is indicated on the right side of the figure.

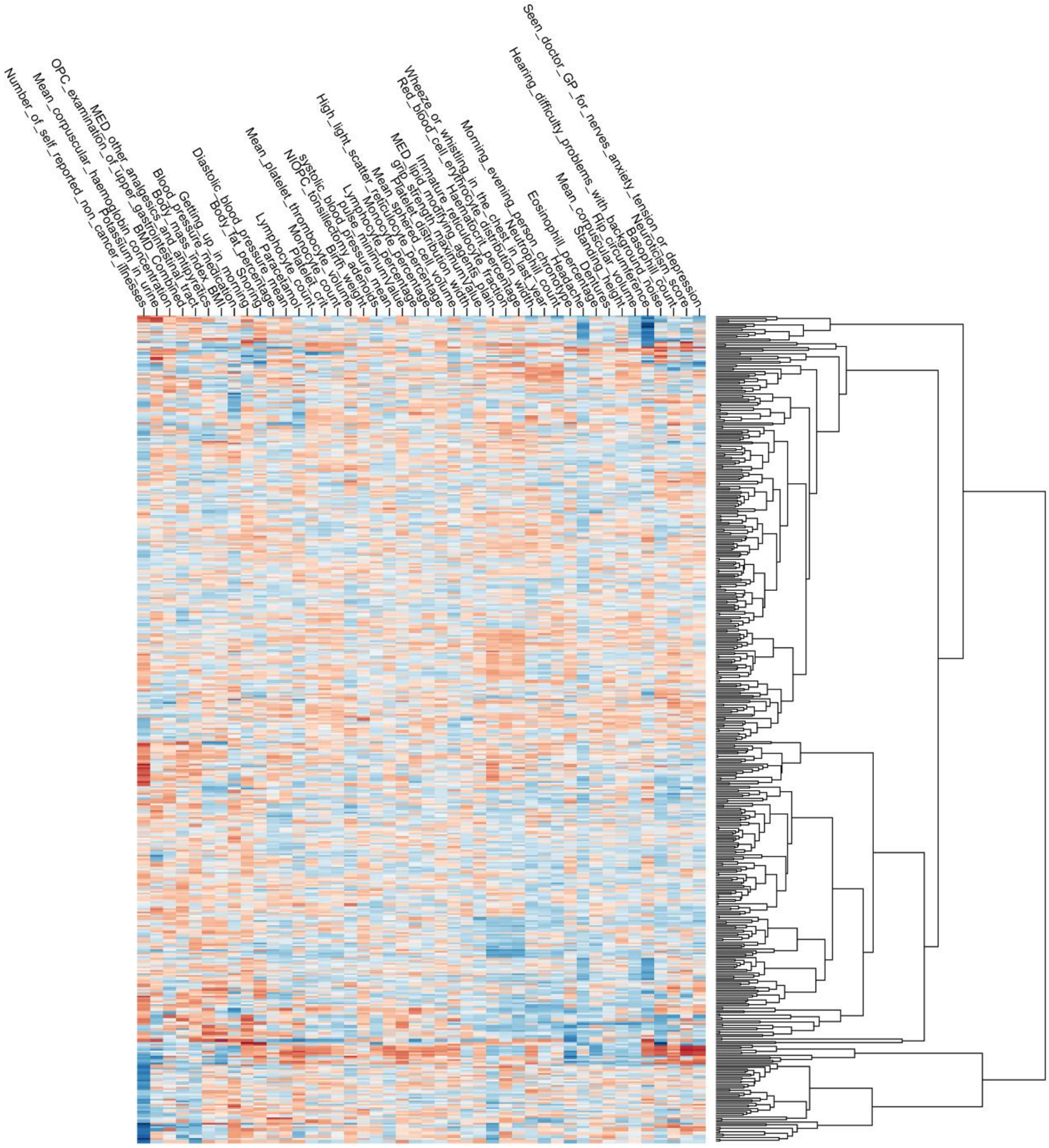

**Figure S3. Alluvial plot illustrating the assignment of variants in the three clusters.**
The alluvial plot provides an overview of the assignment of the 164, 279, and 45 variants for COPD, lung function, and asthma, into the clusters 1, 2 and 3. Note that the three clusters included 156, 148 and 78 variants, respectively, with a subset of 106 variants that could not be formally assigned to any cluster.

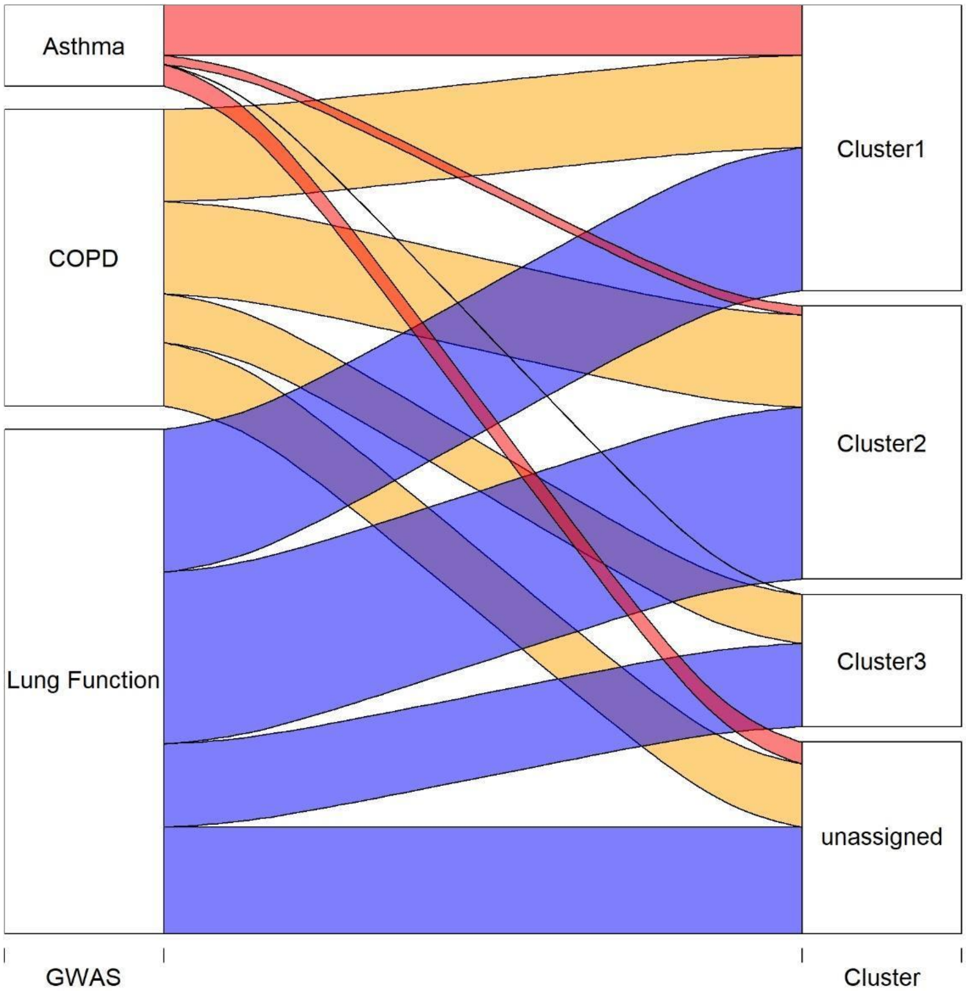

**Figure S4. Marginal effect of genetic risk score across COPDgene phenotypes**

We plotted the association Zscore for the 61 phenotypes for which at least one GRS reaches the Bonferroni corrected p-value threshold of 7 x $10^{-5}$. The red dash lines indicate the significance p-value threshold. Phenotypes are ordered based on the largest absolute Zscore across the four GRS. Phenotypes for which one of $GRS_{1-3}$ was more associated than $GRS_0$ are indicated by a star.

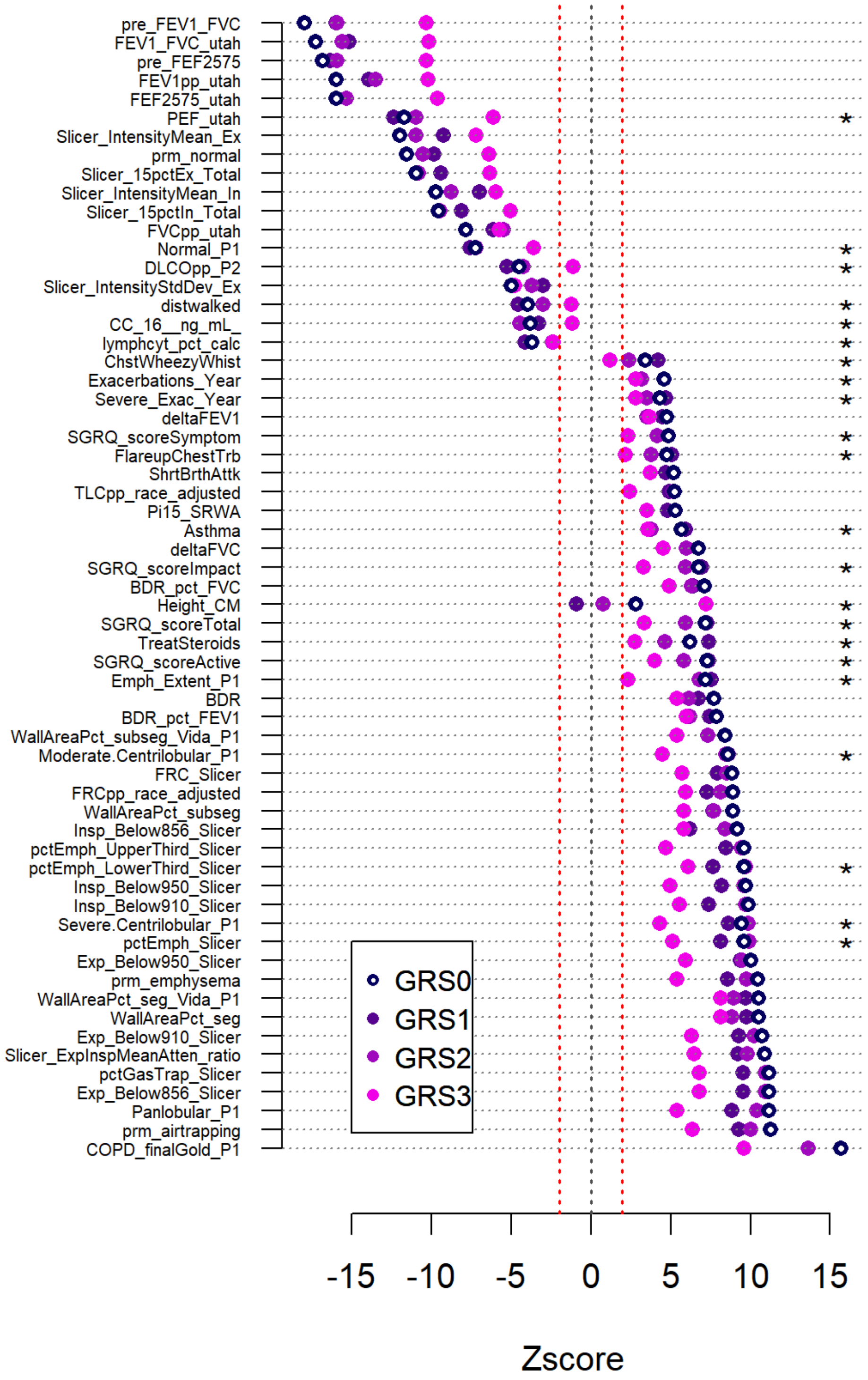

**Figure S5. Frequency of treatments in COPDGene conditional on the genetic risk score**

We derived the frequency of COPD treatments in the whole COPDgene cohort (red bars) and in sub-samples defined based on the three genetic risk scores (GRS). Frequencies in the top 5% percentile of GRS1 (g1+), GRS2 (g2+), and GRS3 (g3+) are indicated by grey bars, and frequencies in the lowest 5% percentiles of GRS1 (g1-), GRS2 (g2-), and GRS3 (g3-) are indicated by orange bars. Final Gold stage (panel top left) was included for comparison purposes.

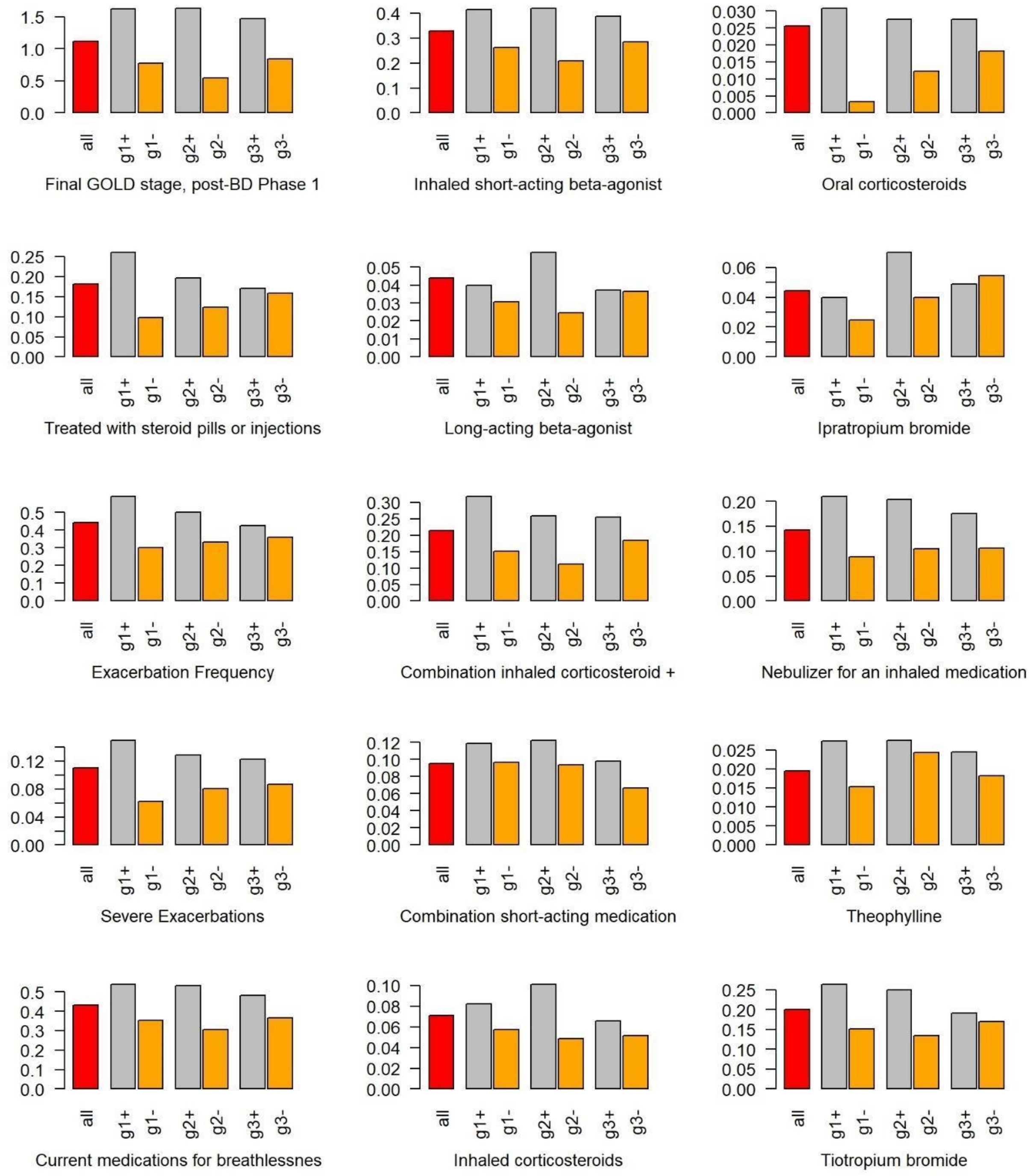

#### COPDGene Acknowledgements:

COPDGene Phase 3
Grant Support and Disclaimer
The project described was supported by Award Number U01 HL089897 and Award Number U01 HL089856 from the National Heart, Lung, and Blood Institute. The content is solely the responsibility of the authors and does not necessarily represent the official views of the National Heart, Lung, and Blood Institute or the National Institutes of Health.
COPD Foundation Funding
COPDGene is also supported by the COPD Foundation through contributions made to an Industry Advisory Board that has included AstraZeneca, Bayer Pharmaceuticals, Boehringer-Ingelheim, Genentech, GlaxoSmithKline, Novartis, Pfizer, and Sunovion.
COPDGene® Investigators – Core Units
Administrative Center: James D. Crapo, MD (PI); Edwin K. Silverman, MD, PhD (PI); Barry J. Make, MD; Elizabeth A. Regan, MD, PhD
Genetic Analysis Center: Terri H. Beaty, PhD; Peter J. Castaldi, MD, MSc; Michael H. Cho, MD, MPH; Dawn L. DeMeo, MD, MPH; Adel El Boueiz, MD, MMSc; Marilyn G. Foreman, MD, MS; Auyon Ghosh, MD; Lystra P. Hayden, MD, MMSc; Craig P. Hersh, MD, MPH; Jacqueline Hetmanski, MS; Brian D. Hobbs, MD, MMSc; John E. Hokanson, MPH, PhD; Wonji Kim, PhD; Nan Laird, PhD; Christoph Lange, PhD; Sharon M. Lutz, PhD; Merry-Lynn McDonald, PhD; Dmitry Prokopenko, PhD; Matthew Moll, MD, MPH; Jarrett Morrow, PhD; Dandi Qiao, PhD; Elizabeth A. Regan, MD, PhD; Aabida Saferali, PhD; Phuwanat Sakornsakolpat, MD; Edwin K. Silverman, MD, PhD; Emily S. Wan, MD; Jeong Yun, MD, MPH
Imaging Center: Juan Pablo Centeno; Jean-Paul Charbonnier, PhD; Harvey O. Coxson, PhD; Craig J. Galban, PhD; MeiLan K. Han, MD, MS; Eric A. Hoffman, Stephen Humphries, PhD; Francine L. Jacobson, MD, MPH; Philip F. Judy, PhD; Ella A. Kazerooni, MD; Alex Kluiber; David A. Lynch, MB; Pietro Nardelli, PhD; John D. Newell, Jr., MD; Aleena Notary; Andrea Oh, MD; Elizabeth A. Regan, MD, PhD; James C. Ross, PhD; Raul San Jose Estepar, PhD; Joyce Schroeder, MD; Jered Sieren; Berend C. Stoel, PhD; Juerg Tschirren, PhD; Edwin Van Beek, MD, PhD; Bram van Ginneken, PhD; Eva van Rikxoort, PhD; Gonzalo Vegas Sanchez-Ferrero, PhD; Lucas Veitel; George R. Washko, MD; Carla G. Wilson, MS;
PFT QA Center, Salt Lake City, UT: Robert Jensen, PhD
Data Coordinating Center and Biostatistics, National Jewish Health, Denver, CO: Douglas Everett, PhD; Jim Crooks, PhD; Katherine Pratte, PhD; Matt Strand, PhD; Carla G. Wilson, MS
Epidemiology Core, University of Colorado Anschutz Medical Campus, Aurora, CO: John E. Hokanson, MPH, PhD; Erin Austin, PhD; Gregory Kinney, MPH, PhD; Sharon M. Lutz, PhD; Kendra A. Young, PhD
Mortality Adjudication Core: Surya P. Bhatt, MD; Jessica Bon, MD; Alejandro A. Diaz, MD, MPH; MeiLan K. Han, MD, MS; Barry Make, MD; Susan Murray, ScD; Elizabeth Regan, MD; Xavier Soler, MD; Carla G. Wilson, MS
Biomarker Core: Russell P. Bowler, MD, PhD; Katerina Kechris, PhD; Farnoush Banaei-Kashani, PhD

COPDGene® Investigators – Clinical Centers

Ann Arbor VA: Jeffrey L. Curtis, MD; Perry G. Pernicano, MD

Baylor College of Medicine, Houston, TX: Nicola Hanania, MD, MS; Mustafa Atik, MD; Aladin Boriek, PhD; Kalpatha Guntupalli, MD; Elizabeth Guy, MD; Amit Parulekar, MD;

Brigham and Women's Hospital, Boston, MA: Dawn L. DeMeo, MD, MPH; Craig Hersh, MD, MPH; Francine L. Jacobson, MD, MPH; George Washko, MD

Columbia University, New York, NY: R. Graham Barr, MD, DrPH; John Austin, MD; Belinda D'Souza, MD; Byron Thomashow, MD

Duke University Medical Center, Durham, NC: Neil MacIntyre, Jr., MD; H. Page McAdams, MD; Lacey Washington, MD

HealthPartners Research Institute, Minneapolis, MN: Charlene McEvoy, MD, MPH; Joseph Tashjian, MD

Johns Hopkins University, Baltimore, MD: Robert Wise, MD; Robert Brown, MD; Nadia N. Hansel, MD, MPH; Karen Horton, MD; Allison Lambert, MD, MHS; Nirupama Putcha, MD, MHS

Lundquist Institute for Biomedical Innovation at Harbor UCLA Medical Center, Torrance, CA: Richard Casaburi, PhD, MD; Alessandra Adami, PhD; Matthew Budoff, MD; Hans Fischer, MD; Janos Porszasz, MD, PhD; Harry Rossiter, PhD; William Stringer, MD

Michael E. DeBakey VAMC, Houston, TX: Amir Sharafkhaneh, MD, PhD; Charlie Lan, DO

Minneapolis VA: Christine Wendt, MD; Brian Bell, MD; Ken M. Kunisaki, MD, MS

Morehouse School of Medicine, Atlanta, GA: Eric L. Flenaugh, MD; Hirut Gebrekristos, PhD; Mario Ponce, MD; Silanath Terpenning, MD; Gloria Westney, MD, MS

National Jewish Health, Denver, CO: Russell Bowler, MD, PhD; David A. Lynch, MB

Reliant Medical Group, Worcester, MA: Richard Rosiello, MD; David Pace, MD

Temple University, Philadelphia, PA: Gerard Criner, MD; David Ciccolella, MD; Francis Cordova, MD; Chandra Dass, MD; Gilbert D'Alonzo, DO; Parag Desai, MD; Michael Jacobs, PharmD; Steven Kelsen, MD, PhD; Victor Kim, MD; A. James Mamary, MD; Nathaniel

Marchetti, DO; Aditi Satti, MD; Kartik Shenoy, MD; Robert M. Steiner, MD; Alex Swift, MD; Irene Swift, MD; Maria Elena Vega-Sanchez, MD

University of Alabama, Birmingham, AL: Mark Dransfield, MD; William Bailey, MD; Surya P. Bhatt, MD; Anand Iyer, MD; Hrudaya Nath, MD; J. Michael Wells, MD

University of California, San Diego, CA: Douglas Conrad, MD; Xavier Soler, MD, PhD; Andrew Yen, MD

University of Iowa, Iowa City, IA: Alejandro P. Comellas, MD; Karin F. Hoth, PhD; John Newell, Jr., MD; Brad Thompson, MD

University of Michigan, Ann Arbor, MI: MeiLan K. Han, MD MS; Ella Kazerooni, MD MS; Wassim Labaki, MD MS; Craig Galban, PhD; Dharshan Vummidi, MD

University of Minnesota, Minneapolis, MN: Joanne Billings, MD; Abbie Begnaud, MD; Tadashi Allen, MD

University of Pittsburgh, Pittsburgh, PA: Frank Sciurba, MD; Jessica Bon, MD; Divay Chandra, MD, MSc; Joel Weissfeld, MD, MPH

University of Texas Health, San Antonio, San Antonio, TX: Antonio Anzueto, MD; Sandra Adams, MD; Diego Maselli-Caceres, MD; Mario E. Ruiz, MD; Harjinder Singh
